# *SLCO1B1* functional variants and statin-induced myopathy in people with recent genealogical ancestors from Africa: a population-based real-world study

**DOI:** 10.1101/2023.12.02.23299324

**Authors:** Sook Wah Yee, Tanushree Haldar, Mark Kvale, Jia Yang, Michael P Douglas, Akinyemi Oni-Orisan

## Abstract

**Background:** Clinical pharmacogenetic implementation guidelines for statin therapy are derived from evidence of primarily Eurocentric study populations. Functional *SLCO1B1* variants that are rare in these study populations have not been investigated as a determinant of statin myotoxicity and are thus missing from guideline inclusion.

**Objective:** Determine the relationship between candidate functional *SLCO1B1* variants and statin-induced myopathy in people with recent genealogical ancestors from Africa.

**Design:** Population-based pharmacogenetic study using real-world evidence from electronic health record-linked biobanks

**Setting:** Various health care settings

**Participants:** Self-identified white and Black statin users with genome-wide genotyping data available.

**Measurements:** Primarily, the odds of statin-induced myopathy + rhabdomyolysis. Secondarily, total bilirubin levels. Thirdly, cell-based functional assay results.

**Results:** Meta-analyses results demonstrated an increased risk of statin-induced myopathy + rhabdomyolysis with c.481+1G>T (odds ratio [OR] = 3.27, 95% confidence interval [CI] 1.43-7.46, *P*=.005) and c.1463G>C (OR = 2.45, 95% CI 1.04-5.78, *P*=.04) for Black participants. For White participants, c.521T>C was also significantly associated with increased risk of statin-induced myopathy + rhabdomyolysis (OR = 1.41, 95% CI 1.20-1.67, *P*=5.4x10^−5^). This effect size for c.521T>C was similar in the Black participants, but did not meet the level of statistical significance (OR = 1.47, 95% CI 0.58-3.73, *P*=0.41). Supporting evidence using total bilirubin as an endogenous biomarker of *SLCO1B1* function as well as from cell-based functional studies corroborated these findings.

**Limitations:** Data limited to severe statin myotoxicity events.

**Conclusion:** Our findings implicate Afrocentric *SLCO1B1* variants on preemptive pharmacogenetic testing panels, which could have an instant impact on reducing the risk of statin-associated myotoxicity in historically excluded groups.

**Primary Funding Source:** National Institutes of Health, Office of the Director - All of Us (OD-AoURP)

## Introduction

3-Hydroxy-3-methylglutaryl coenzyme A (HMG-CoA) reductase inhibitors (more commonly known as statins) are among the most efficacious therapies for the prevention of atherosclerotic cardiovascular disease (ASCVD); however, their broader population-wide utilization is substantially hampered from the adverse drug reaction of myotoxicity(1). Pharmacogenetic findings linking elevated statin drug levels to myotoxicity suggest that these debilitating adverse drug reactions cannot be entirely attributed to a nocebo affect and that a precision medicine approach to statin selection may improve outcomes(2).

Currently, the Clinical Pharmacogenetics Implementation Consortium (CPIC) provides recommendations with the highest strength of evidence for statin prescribing based on *SLCO1B1* genetic test results (*SLCO1B1* encodes the liver-specific organic anion transporting polypeptide [OATP1B1]) to reduce the risk of myotoxicity(3). Consequently, statin pharmacogenetics is among the most common clinical pharmacogenetic implementation examples in practice(4); the inclusion of *SLCO1B1* as part of a 12-gene pharmacogenetic test panel in a landmark randomized clinical trial was a major contributor to the primary outcome showing a 30% relative risk reduction for general adverse drug reactions(5). However, the evidence for which CPIC recommendations of statins and other therapies derive from are based on largely Eurocentric study populations (i.e., participants who predominantly self-identify as White and/or derive recent ancestry from any region within the continent of Europe)(2,6). As a result, variants known to impact OATP1B1 function that are rare in Eurocentric but common in non-Eurocentric groups (i.e., one or more of the diverse groups that do not fit the above definition of Eurocentric) have not been investigated as a determinant of statin myotoxicity and are thus missing from guideline inclusion.

Taken together, these findings demonstrate that pharmacogenetic tests for statin therapy are not providing equitable benefit in preventing the risk of adverse drug reactions. Enhanced investigation in non-Eurocentric groups will generate the evidence base necessary for more inclusive pharmacogenetic prescribing guidelines. The objective of this study was to determine the relationship between candidate functional *SLCO1B1* variants and statin-induced myopathy in people with recent genealogical ancestors from Africa.

## Methods

### Data Resources

We conducted our studies using de-identified, individual-level, population-based data capturing the health of participants from three independent electronic health record-linked biobanks (each with genome-wide genotype data): the National Institutes of Health (NIH) All Of Us research program (AoURP), the Genetic Epidemiology Research on Adult Health and Aging (GERA) Cohort, and the UK Biobank (UKB). Institutional Review Board (IRB) approval was obtained from both Kaiser Permanente (Mid-Atlantic States Region) and the University of California. Participants gave written informed consent. See Supplementary Material for further details.

### Dataset Types

Within each data resource, we used electronic health record, self-administered survey, physical measurement, and genomic data. Further details are available in the Supplementary Material.

### Justification for the use of population descriptors in genetic studies

Participants who self-identified as white or Black were included in the study. The selection of these descriptors were carefully chosen and in concordance with guidance from the National Academies of Sciences, Engineering, and Medicine(7). It was critical to have these groups in order to showcase the disparity, as previously done in similar studies for health disparities precision medicine research(8). Further details are available in the Discussion as well as the Supplementary Material. The specific definition for these race categories in each cohort are described in the Data Resources section of the Supplementary Material.

### Genotype

Consistent with the objective of this study, we selected candidate *SLCO1B1* variants with evidence of mechanistic function that are common (> 1% allele frequency) in at least one of the many groups within the African (AFR) superpopulation of the 1000 Genomes data set (available through the online gnomAD browser: https://gnomad.broadinstitute.org/). A systematic search identified 3 variants that met these criteria: missense variant reference single nucleotide polymorphism(rs)4149056 (c.521T>C, OATP1B1-Val174Ala), missense variant rs59502379 (c.1463G>C, OATP1B1-Gly488Ala), and loss-of-function splice variant rs77271279 (c.481+1G>T). Each of them happened to have evidence for reduced OATP1B1 function. Further details are available in the Supplementary Material.

### Phenotype (statin-induced myotoxicity)

We generated a phenotype for severe statin-induced myotoxicity, as previously described(9) with modifications. Further details are available in the Methods section and Supplemental Table 1 of the Supplementary Material.

### Phenotype (bilirubin levels)

As a secondary outcome to support our primary objective, we used total bilirubin levels, an endogenous biomarker for OATP1B1 activity(10). Further details are available in the Supplementary Material.

### Cell-based assay studies

We conducted *in vitro* mechanistic studies to support our population-based studies and extend the current functional evidence for our *SLCO1B1* variants of interest. See Supplementary Material for details.

### Statistical Analysis

We determined the relationship between our candidate *SLCO1B1* variants (c.521T>C, c.481+1G>T, c.1463G>C) and statin-induced myopathy + rhabdomyolysis using multiple logistic regression in race-stratified groups from both cohorts with drug response phenotypes (AoURP, GERA). Participant age and self-identified sex at birth were included as covariates. Our primary analyses were meta-analyses across cohorts using a fixed-effects model of the relationship within each race strata for all statin types combined. For our secondary analyses, we investigated the association between each of our variants and total bilirubin levels in self-identified Black participants (UKB, AoURP). To do this, we performed a linear regression model on log_10_-transformed total bilirubin levels. Age, sex, body mass index, and 10 principal components of AFR-specific genetic similarity were included as covariates. We also conducted a sensitivity analysis by limiting the study population to the subset with total bilirubin levels within the range of normal (1.71-20.50 micromoles per liter for UKB and 0.1-1.2 milligrams per deciliter for AoURP). All analyses were conducted using R software (version 4.3.1; R Foundation for Statistical Computing; www.R-project.org; Vienna, Austria). Regression models were constructed using the ‘glm’ function in R. We used the metafor R package for our meta-analyses. STROBE guidelines were followed for reporting results. Differences in transporter uptake and cytotoxicity for our cell-based studies were tested by one-way analysis of variance (ANOVA) followed by Tukey’s multiple comparison tests. *P* values < 0.05 were considered statistically significant.

## Results

### Characterization of the study population

A total of 555 statin-induced myopathy + rhabdomyolysis cases (200 myopathy + 355 rhabdomyolysis) met the criteria of study inclusion for our primary analysis. Among these, 71 identified as Black and 484 identified as white. Low allele frequencies for c.1463G>C and c.481+1G>T in white participants precluded subsequent analyses for those variants in this subset of the study population. Supplemental Table 2 provides more details for this population.

### The relationship between *SLCO1B1* variants and statin-induced myopathy + rhabdomyolysis

Our meta-analyses results demonstrated an increased risk of statin-induced myopathy + rhabdomyolysis with c.481+1G>T (odds ratio [OR] = 3.27, 95% confidence interval [CI] 1.43-7.46, *P*=.005) and c.1463G>C (OR = 2.45, 95% CI 1.04-5.78, *P*=.04) for Black participants (Table 1). For White participants, c.521T>C was also significantly associated with increased risk of statin-induced myopathy + rhabdomyolysis (OR = 1.41, 95% CI 1.20-1.67, *P*=5.4x10^−5^). This effect size for c.521T>C was similar in the Black participants, but did not meet the level of statistical significance (OR = 1.47, 95% CI 0.58-3.73, *P*=0.41). Cohort-specific results are displayed in Table 1.

**Table 1.** Relationship1 between candidate functional *SLCO1B1* variants and statin-induced myopathy + rhabdomyolysis.

| *SLCO1B1* variant | Analysis group<sup>2</sup> | Cohort | n (case/control) | Beta | SE | Odds Ratio | 95% CI | P-value |
| --- | --- | --- | --- | --- | --- | --- | --- | --- |
| c.481+1G>T | Black | Meta-analysis | 71/702 | 1.18 | 0.42 | 3.27 | 1.43-7.46 | 0.0049 |
|  |  | AoURP | 31/310 | 1.17 | 0.52 |  |  |  |
|  |  | GERA | 40/392 | 1.20 | 0.70 |  |  |  |
| c.1463G>C | Black | Meta-analysis | 71/702 | 0.90 | 0.44 | 2.45 | 1.04-5.78 | 0.040 |
|  |  | AoURP | 31/310 | 1.19 | 0.46 |  |  |  |
|  |  | GERA | 40/392 | -1.90 | 1.41 |  |  |  |
| c.521T>C | Black | Meta-analysis | 71/702 | 0.39 | 0.47 | 1.47 | 0.58-3.73 | 0.41 |
|  |  | AoURP | 31/310 | -0.88 | 1.04 |  |  |  |
|  |  | GERA | 40/392 | 0.72 | 0.53 |  |  |  |
| c.521T>C | White | Meta-analysis | 484/4,840 | 0.35 | 0.09 | 1.41 | 1.20-1.67 | 5.35x10<sup>-5</sup> |
|  |  | AoURP | 38/380 | 0.40 | 0.32 |  |  |  |
|  |  | GERA | 446/4,460 | 0.34 | 0.09 |  |  |  |

<sup>1</sup>Generated by logistic regression. Covariates included age and sex at birth. Meta-analyses used a fixed-effects model.
<sup>2</sup>Self-identified race
AoURP, All of Us Research Program; CI, confidence interval; GERA, Genetic Epidemiology Research on Adult Health and Aging; SE, standard error

### The relationship between *SLCO1B1* variants and total bilirubin levels

A total of 29,119 self-identified Black participants from UKB met the study inclusion criteria for the secondary analysis (6,651 from UKB and 22,468 from AoURP). Baseline characteristics in this total bilirubin study population are shown in Supplemental Table 3. All three variants of interest were associated with elevated total bilirubin levels. The strongest association was observed for c.481+1G>T (beta = 0.13, p=1.1x10^−9^) followed by c.1463G>C (beta = 0.08, p=6.5x10^−6^) and c.521T>C (beta=0.05, p=0.024). Consistent results were observed from AoURP (Supplemental Table 4). Sensitivity analyses also showed a similar association (Supplemental Table 5).

### Cell-based mechanistic studies for OATP1B1-Val174Ala and OATP1B1-Gly488Ala

Our HEK293T landing cells transiently overexpressing OATP1B1 reference, OATP1B1-Val174Ala, and OATP1B1-Gly488Ala showed significantly greater green fluorescent protein (GFP) abundance compared to HEK293T landing cells without GFP expression (Figure 1A). Additionally, the OATP1B1-Val174Ala and OATP1B1-Gly488Ala missense variants exhibited similar GFP abundance when compared to OATP1B1 reference (Figure 1A). Moreover, OATP1B1 reference, OATP1B1-Val174Ala, and OATP1B1-Gly488Ala each had plasma membrane GFP expression (Figure 1B).

**Figure 1.**
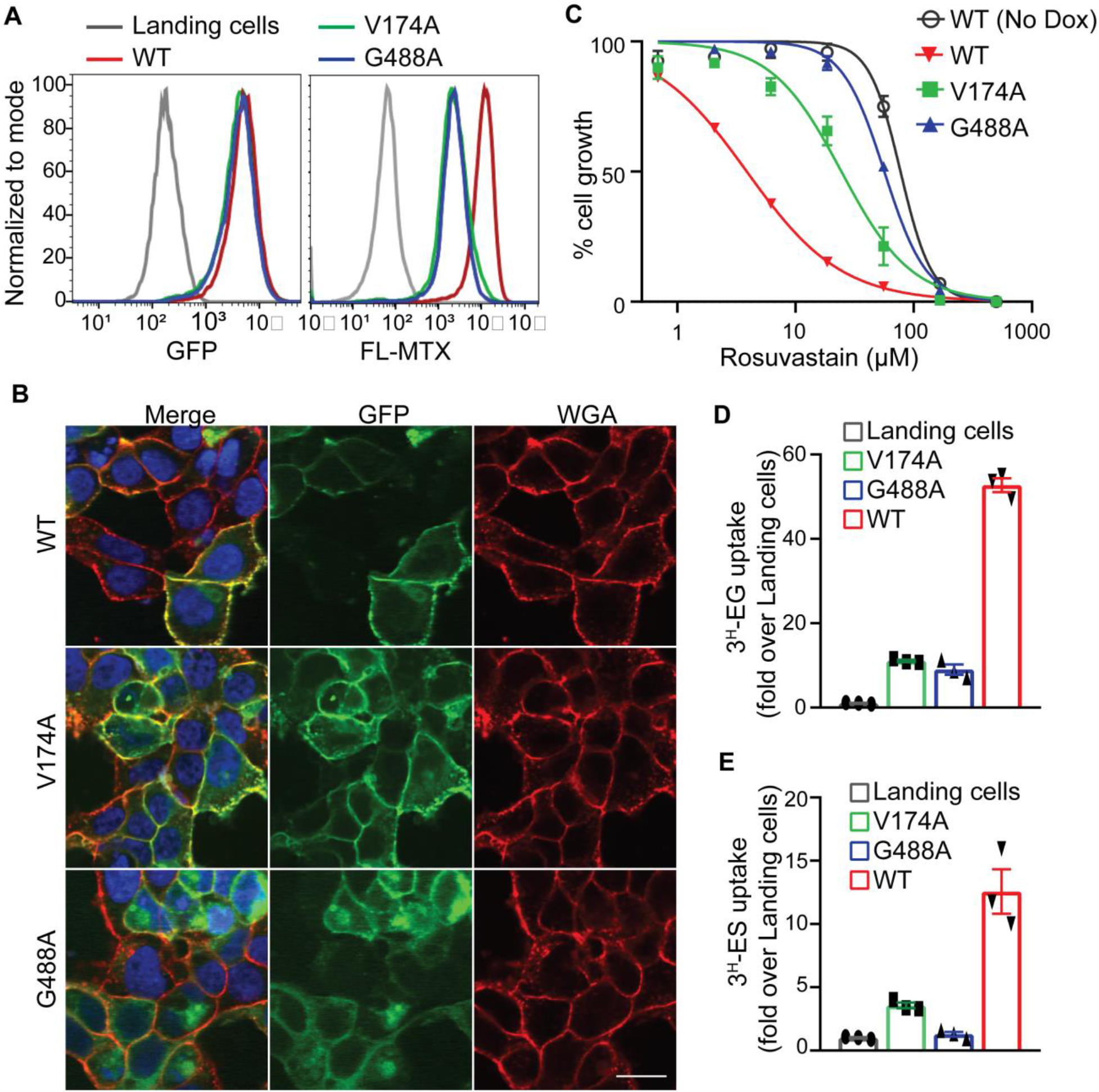
Functional characterizations of two missense variants in OATP1B1. (A) OATP1B1 reference (WT) and the two missense OATP1B1 variants (V174A and G488A) had similar green fluorescent protein (GFP) expression as well as levels greater than that of HEK293T landing cells without GFP (left). OATP1B1 WT has increased uptake of OATP1B1 substrate fluoresceine-methotrexate (FL-MTX) compared to the missense variants (right). (B) Cellular localization of GFP-tagged OATP1B1 WT and V174A has overlap with the plasma membrane marker wheat germ agglutinin (WGA; red). OATP1B1 G488A showed weaker expression on the plasma membrane and some intracellular localization. (C) Cytotoxicity of rosuvastatin was greatest in HEK293T landing pad cells stably transfected with OATP1B1 WT (red), followed by OATP1B1-V174A (green) then OATP1B1-G488A (blue) compared to cells without OATP1B1 (open circle). Both missense OATP1B1 variants ((V174A and G488A) showed significantly reduced uptake of radioligands (D) [3H]-estradiol-17beta-glucuronide (EG) and (E) [3H]-esterone sulfate (ES) compared to OATP1B1 WT.

Despite similar GFP expression compared to OATP1B1 reference, the missense variants demonstrated significantly reduced intracellular fluorescence as well as reduced uptake of both estrone sulfate and estradiol-17β-glucuronide (Figures 1A, 1D, 1E). Furthermore, only OATP1B1-Gly488Ala showed mixed localization of GFP abundance on the plasma membrane and intracellularly (Figure 1B). There was a slight reduction of OATP1B1-Gly488Ala on the plasma membrane compared to OATP1B1-Val174Ala. Accordingly, OATP1B1-Gly488Ala cells showed a reduced uptake of estrone-sulfate and less cytotoxicity to rosuvastatin (Figure 1C and 1E) compared to OATP1B1-Val174Ala. However, the uptake of estradiol-17β-glucuronide was not significantly different between OATP1B1-Val174Ala and OATP1B1-Gly488Ala (Figure 1D).

## Discussion

This investigation is the first to demonstrate the clinical validity for statin-induced myopathy of variants that are common in genealogical ancestors from Africa. Using electronic health record-linked biobanks, we generated the largest cohort of statin-induced myopathy in study populations of self-identified Black participants. Using this cohort and matched controls, we found that two functional *SLCO1B1* variants extremely rare in Eurocentric groups were strongly associated with this adverse drug reaction, an effect of greater magnitude than our results for the well-established c.521T>C variant within any group. We reinforced the importance of these Afrocentric (AFR-specific) variants using clinical studies for an endogenous biomarker of OATP1B1 as well as cell-based functional studies. Considering the magnitude of effect for these variants and the unprecedented size of the study population for our primary statin response analysis as well as our strong supporting evidence, our translational findings provide strong rationale for the inclusion of these variants on pharmacogenetic testing panels.

Adverse drug reactions are a common, underappreciated cause of morbidity and mortality; two million hospitalizations each year in the United States are caused by these events according to the Department of Health and Human Services. Pharmacogenetics is widely considered an effective tool to combat some adverse drug reactions, especially for events unrelated to prescribing errors (e.g., adverse drug reactions that occur despite a drug being taken at the recommended normal dose). Considering that 1) the vast majority of patients undergoing routine care (>90%) carry at least one pharmacogenetic variant that would warrant a change in therapy to avoid an adverse drug reaction(11) and that 2) preemptive pharmacogenetic prescribing has demonstrated efficacy in reducing the relative risk of adverse drug reactions by almost one third(5), the universal implementation of this precision medicine approach to pharmacotherapeutic prescribing shows great promise in improving health outcomes on a large scale. As a consequence, preemptive genotyping, which allows clinicians to have the genetic data at the time of drug prescribing, is gaining wider acceptance and increased coverage by insurance plans(12,13).

However, there is also a growing concern among clinicians and scientists that, if not done carefully, the implementation of precision medicine tools has the potential to exacerbate preexisting health disparities(14). In particular, taking results from Eurocentric study populations and applying them to non-Eurocentric groups (without considering that average results vary between stratified groups) has the potential to cause harm(15). Conducting analyses in genetically diverse study populations and considering all the factors that impact disparities are proposed solutions to prevent further exacerbation(16).

For example, one study found that using a normal range for absolute neutrophil count (“normal” is based on results from Eurocentric study populations) to guide azathioprine immunosuppressant therapy (a standard of care practice in United States health systems) leads to disproportionately higher rates of unnecessary therapeutic discontinuation in Black patients. Even after adjusting for non-genetic factors, this disparity was attributed entirely to an *ACKR1* variant (present in > 80% of Black and <0.5% of white participants, gnomad.broadinstitute.org), a variant associated with low (relative to “normal”) but safe neutrophil counts(8). Findings suggest that pharmacogenetic testing for *ACKR1* in all patients to guide azathioprine therapy may prevent the exacerbation of this disparity, which already exists for reasons unrelated to pharmacogenetics(17). Importantly, this potential exacerbation would never have been discovered if the investigators did not stratify groups with the use of population descriptors (90% of the study population was White, which would have diluted results in a pooled analysis). Similarly, another study found that a Eurocentric precision medicine tool for warfarin dosing led to less accurate anti-coagulation in the subset of the study population who were Black(18). Accounting for pharmacogenetic variants not commonly found in Eurocentric study populations improved dosing for Black participants(19,20). These are just a couple of many other examples (21–24) demonstrating the usefulness of population descriptors to identify potential precision-medicine relevant health disparity exacerbations that otherwise could never be discovered.

The potential for exacerbation of existing health disparities from precision medicine is no different for statin therapy. It’s already known that racial disparities in the use of statin therapy exist, an observation thought to be attributable to implicit racism(25). Thus, it would be important that statin precision medicine tools designed to prevent myotoxicity and thereby promote continued therapeutic use would work equitably across multiple groups of people. This is especially true considering that statins are consistently among the most commonly chosen medication classes included in clinical pharmacogenetic implementation programs(5,12) and are among the most “actionable” (i.e., patients taking statins are carriers of genotypes for which guidelines would suggest a therapeutic regimen change)(5). CPIC guidelines recommend (with level A evidence: the highest classification rating) specific statin types and doses for people with OATP1B1 decreased or poor function phenotype for who the drug is indicated(3). Presumably, considering the strength of the recommendation, this pharmacogenetic test should benefit everyone. However, the guidelines dictate that the OATP1B1 phenotype should be determined only by haplotypes containing c.521T>C (e.g.,*\*5* and *\*15*), which have lower allele frequency in people with recent genealogical ancestors from Africa (<0.5% prevalence in multiple groups within the African superpopulation including ESN, YRI, GWD, and MSL) including self-identified Black Americans. This suggests that the currently recommended *SLCO1B1*-based pharmacogenetic test may be less beneficial in people with recent genealogical ancestors from Africa. Our results replicated these allele frequencies and validated the hypothesis that this pharmacogenetic test does not benefit groups equitably on a population-wide scale.

In contrast to c.521T>C, there are *SLCO1B1* variants that are rare in Eurocentric study populations. In particular, two *SLCO1B1* polymorphisms (missense variant c.1463G>C and splice donor c.481+1G>T) are each independently associated with levels of endogenous OATP1B1 substrates and have mechanistic evidence for reduced transport function(26–29). These variants were discovered in Afrocentric study populations and are only common (>1% allele frequency) in people with recent genealogical ancestors from Africa (e.g., subpopulations from multiple regions within and outside of Africa including ASW, ACB, LWK, ESN, YRI, GWD, and MSL). They have been entirely understudied as pharmacogenetic markers.

We sought to provide clinical validity for these understudied variants in the context of statin-induced myopathy. We found that these variants had a strong association with severe statin-induced myotoxicity across all statin types. Notably, the clinical significance of this findings is substantial. Our reported effect sizes were stronger than what we found for the c.521T>C association with statin-induced myopathy in our Eurocentric study populations. Between the two AFR-specific variants, c.481+1G>T was the strongest; this observation is not inconsistent with the large effects observed by loss of function variants relative to missense polymorphisms. Our total bilirubin and cell-based studies support these relative effect sizes. It is also important to remark that these AFR-specific variants may have been considered too rare (<0.1%) to include if our analyses had been conducted in a pooled study population without the use of descriptors (i.e., not stratified by groups). Altogether, our findings provide evidence for the potential of these variants to prevent statin-induced myopathy events that the standard *SLCO1B1*-based pharmacogenetic test are not able to capture.

Our findings are consistent with a case study reporting atorvastatin-induced rhabdomyolysis in a “51-year-old African American male”(30). The patient carried no copies of the well-established c.521T>C polymorphism. However, he was found to carry one copy of c.481+1G>T, which was thought to be the causative factor of this adverse drug reaction. This patient was ultimately switched to a non-statin lipid lowering alternative. This clinical case further illustrates the potential of these novel pharmacogenetic variants as a preemptive test to prevent myotoxicity before statin initiation.

This case study as well as the current investigation highlights more severe statin reactions across the full spectrum of statin-induced myotoxicities. It is important to note that, even if they don’t lead to hospitalization, less severe statin-induced myotoxicity can still hamper daily activities of living(31) and increase the risk of major adverse cardiovascular outcomes (by promoting statin nonadherence)(32).

Considering the demonstrated association between c.521T>C and these less severe adverse drug reactions as well as for other drugs and toxicities(33–35), the Afrocentric *SLCO1B1* variants likely would prevent these as well. However, additional studies are needed to validate this.

Altogether, our findings provide strong rationale for writing committees to recommend incorporation of Afrocentric *SLCO1B1* variants into clinical pharmacogenetic guidelines. However, considering the clinical significance of these findings and the potential immediate impact on health equity, a strong consideration (at least by health systems with established pharmacogenetic testing programs in place) should be made to implement these variants even sooner than guideline incorporation. Statin-based pharmacogenetic guidelines from CPIC, for example, are released only once every several years and were just recently released as of the writing of this paper(3). Furthermore, our combined 71 cases of severe statin-induced myotoxicity within Black participants is a similar sample size to that from the landmark study (in a Eurocentric study population)(2) for which the original CPIC recommendations(36) are based.

In summary, our findings implicate Afrocentric *SLCO1B1* variants on preemptive pharmacogenetic testing panels, which could have an instant impact on the risk of statin-associated myotoxicity. Most importantly, these findings set the foundation for future studies that deviate from the sole reliance on Eurocentric study populations to improve pharmacological health outcomes in all.

## Data Availability

All summary-level data produced in the present study are available upon reasonable request to the authors

## Acknowledgement

Research reported in this publication was supported by the National Human Genome Research Institute (R01HG012824: T.H., M.P.D., A.O.) and the National Institute of General Medical Sciences (R01GM117163: S.W.Y.; R01GM139875: K.M.G.) of the NIH as well as the OD-AoURP (which also sponsored R01HG012824: T.H., M.P.D., A.O.). The content is solely the responsibility of the authors and does not necessarily represent the official views of the National Institutes of Health or the OD-AoURP. This study was made possible through data provided by the Kaiser Permanente Division of Research Program on Genes, Environment, and Health (RPGEH) funded by the National Institute on Aging, the National Institute of Mental Health, the National Institute of Health Office of the Director, the Robert Wood Johnson Foundation, and the Kaiser Permanente Community Benefits Program. We would like to express our gratitude to the members of the All of Us Evenings with Genetics Research Team at Baylor College of Medicine, particularly Drs. J. Wang and S.E. Scherer, for their valuable contributions and support throughout this research endeavor.

