## Supplementary Material for "*SLCO1B1* functional variants and statin-induced myopathy in people with recent genealogical ancestors from Africa: a population-based real-world study"

**Methods**

Data Resources.

The National Institutes of Health (NIH) All Of Us research program (AoURP) is an open-access data resource of adults 18 years and older residing in the United States. The AoURP data that we accessed was constructed based on the All of Us Controlled Tier Dataset v7 (curated version 2022Q4R9), which includes data from participants who enrolled between 2018 and 2021. The data goes back as far as 1980 with a data cutoff date of July 1, 2022 (and was released to investigators in Spring 2023). These data were accessed from the Controlled Tier dataset (the only tier with access to genetic data). We analyzed data through the AoURP Researcher Workbench cloud-based platform using AoURP Dataset Builder and Cohort Builder (based on SQL). As of the time of this study, ~413,000 participants had enrolled. Among enrolled participants, there were >280,000 participants with adequate electronic health record, survey, and genomic data from which a study population was selected. Of principal relevance to this study, 78,504 participants self-identified as "Black, African American or African (For example: African American, Ethiopian, Haitian, Jamaican, Nigerian, Somali, etc.)". AoURP data are publicly available by request from https://www.researchallofus.org.

The Genetic Epidemiology Research on Adult Health and Aging (GERA) Cohort is nested within the Research Program on Genes, Environment and Health (RPGEH), which is a large (~400,000) data resource for epidemiological research of common age-related diseases(1). RPGEH participants were recruited from members of Kaiser Permanente Northern California (KPNC), a medical care plan providing comprehensive coverage with the exception of dental and vision. Recruitment took place in 2007 through a mailed survey (capturing demographic, behavioral, and health characteristics). Participation required KPNC membership for ≥2 years, survey completion, a saliva sample for future genetic testing, and consent for access to the participant's electronic health records. GERA is the genetic subset or RPGEH for whom deoxyribonucleic acid (DNA) was extracted from the saliva samples for the generation of genome-wide genotyping data. The data for this study covered records from 1996 to 2022. GERA contains a total of 110,266 participants. Among these, 3,365 participants self-identified as "African American", "African", "Afro-Caribbean" (~93% identified as "African American"). GERA data are accessible through the application and review process of the Kaiser Permanente Research Bank (https://researchbank.kaiserpermanente.org/our-research/for-researchers/).

The UK Biobank (UKB) is an open-access data resource containing adults residing in the United Kingdom (England, Wales, and Scotland)(2). Participants were recruited from the United Kingdom's National Health Service. Recruitment took place between 2006 and 2010 through mailed invitations to individuals within 25 miles of an assessment center. Participants were required to be 40-69 years of age during the time range of recruitment. During the first assessment visit, participants filled out questionnaires, were measured for physical characteristics, and provided biological samples (blood, urine, and saliva). Whole blood samples and serum derived from these samples were tested to generate levels for commonly measured laboratory biomarkers. Participants also provided consent for investigator access to their electronic health records. For this study, UKB data was analyzed with investigators having Tier 2 access under approved application number 14105 (PI: J.S.W.) and in accordance with the UK Biobank Ethics and Governance Framework. The cohort contains 500,000 participants including 8,066 participants who identify as “Black or Black British”. UKB data are publicly available by request from https://www.ukbiobank.ac.uk.

Dataset Types.

Within each data resource, we used electronic health record, self-administered survey, physical measurement, and genomic data.

Electronic health records from at least one of the above data resources included pharmacy records, International Classification of Diseases (ICD) diagnostic codes (both ICD-9 and ICD-10 codes), laboratory measurements, and survey data. Pharmacy records included statins (simvastatin, lovastatin, atorvastatin, pravastatin, and rosuvastatin) and drugs that interact with statins (amprenavir, atazanavir, cyclosporine, darunavir, dasabuvir, lopinavir, fenofibrate, gemfibrozil, indinavir, nelfinavir, ritonavir, saquinavir, tipranavir, amiodarone, verapamil, cimetidine, clarithromycin, cobicistat , diltiazem, dronedarone, erythromycin, fluconazole, fosamprenavir, isavuconazonium, itraconazole, ketoconazole, posaconazole, and voriconazole). ICD codes included rhabdomyolysis (ICD-9 code 728.88, ICD-10 M62.82) and acute myocardial infarction (ICD-9 410.xx, ICD-10 I21.xx, and ICD-10 I22.xx). Lab values included total bilirubin (AoURP) and creatine kinase (CK).

For our genetic data, we used microarray genotyping results available from our data resources (AoURP, GERA, UKB). Only participants with genotyping array data were included in the study.

Other types of data (e.g., survey data, physical measurements, per protocol laboratory biomarkers outside of routine care) were the sources for total bilirubin (UKB), body mass index, self-identified race, date of birth, and self-identified sex at birth.

Justification for the use of population descriptors in genetic studies.

We have carefully considered the available options for the use of population descriptors and consulted the most recent guidance document on this topic (3). Our investigation meets the NIH National Institute on Minority Health and Health Disparities (NIMHD) definition of health disparities research because we report “a health difference, on the basis of one or more health outcomes, that adversely affects disadvantaged populations”(4) and are intended to ultimately improve health outcomes for people who are understudied in pharmacogenetics. Currently, Eurocentric study populations predominate pharmacogenetic findings in the literature, which has the potential to exacerbate preexisting inequity for numerous health conditions(5). For this reason, it was critical for us to include both race and genetic similarity in this study including race-stratified analyses. This is consistent with the recommendation from the most recent guidance document to consider race descriptors in studies that fall under the “Health Disparities with Genomic Data” category and with the wording “In the interests of equity, justice, or the diversification of human genetic data and knowledge about it, researchers may choose to use race and/or ethnicity in order to identify individuals to be included in their studies (Oni-Orisan et al., 2021)” (3,6). Of note, race was not used to control for genetics or as a proxy for genetic similarity. Language throughout this paper was chosen prudently to avoid confusion, assumptions, and misconceptions about human groupings as well as to facilitate replication in future studies. Searches for “Eurocentrism” and “pharmacogenetics” (to gather information on this topic catered to our specific subfield within genetics that could further guide us) from the most recent recommendations(3) yielded no additional aid. Further details on the importance of our selected population descriptors are available in the Discussion section.

Genotype.

Consistent with the objective of this study, we selected candidate *SLCO1B1* variants with evidence of mechanistic function that are common (> 1% allele frequency) in at least one of the many groups within the African (AFR) superpopulation of the 1000 Genomes data set (i.e., African Ancestry in South West USA [ASW], African Caribbean in Barbados [ACB], Esan in Nigeria [ESN], Gambian in Western Division – Mandinka [GWD], Luhya in Webuye, Kenya [LWK], Mende in Sierra Leone [MSL], Yoruba in Ibadan, Nigeria [YRI]). A comprehensive search was conducted within the online gnomAD browser (https://gnomad.broadinstitute.org/) and PubMed database. Two independent investigators (S.W.Y. and A.O.) conducted the search using relevant genetic terms ("SLCO1B1" or "OATP1B1") in combination with relevant populations descriptors ("sub-Saharan", "African", "Black", "AFR", or "Afrocentric"). We identified 3 variants that met these criteria: missense variant reference single nucleotide polymorphism(rs)4149056 (c.521T>C, OATP1B1-Val174Ala), missense variant rs59502379 (c.1463G>C, OATP1B1-Gly488Ala), and loss-of-function splice variant rs77271279 (c.481+1G>T)(7–12). Each of them happened to have evidence for reduced OATP1B1 function. Both c.1463G>C and c.481+1G>T were common in each of the 1000 Genomes groups within superpopulation AFR, but in no other group on a global scale. Only c.521T>C was common in ≥ 1 group outside of AFR. This variant was only common in 3 of the 7 AFR groups (ACB, ASW, LWK) with a much lower allele frequency compared to all other superpopulations (e.g., 0.015 in AFR versus 0.161 in EUR).

Genetic similarity eigenvectors previously generated from principal component analyses of the microarray genotyping data were also used(7). These genetic similarity factors were used as covariates. Slight variations in the methods between both cohorts are presented in Supplemental Table 1.

Phenotype (statin-induced myotoxicity).

We generated a phenotype for severe statin-induced myotoxicity, as previously described (8)with modifications as follows.

Severe statin-induced myotoxicity events were classified into two types: statin-induced myopathy and statin-induced rhabdomyolysis.

A participant met the criteria for a statin-induced myopathy case with ≥ 1 qualifying CK level > 5× the upper limit of normal (ULN) within the timeframe of a high-dose statin pharmacy record (between the start date of the pharmacy record and 7 days following the end date of that record). We set the ULN for CK as 336 units per liter for males and 176 units per liter for females. High-dose statin therapy was defined as ≥ 40 milligrams daily regardless of statin type. We used two criteria to qualify a CK level. First, myopathy events recorded within 7 days of a myocardial infarction, the most common cause of elevated CK(9), were not considered as an eligible event that could qualify a participant as a case. Second, we also disqualified events of which there was ≥ 1 pharmacy record for drugs that strongly interact with statins (if within a half year prior to event date). See Dataset Types section for the list of interacting drugs.

Statin-induced rhabdomyolysis cases were defined as participants with ≥ 1 qualifying diagnosis of rhabdomyolysis event within 7 days after receiving a high-dose statin prescription. We disqualified rhabdomyolysis events with evidence of a drug interaction as above (i.e., ≥ 1 pharmacy record for drugs that interact with statins if within a half year prior to event date).

For participants with multiple qualifying myotoxicity events, only one event was used for each participant. In this situation, the earliest event was chosen as the index event, the statin filled most immediately before the index event was deemed to be the index statin, and the start date for the index statin was considered the index date. The only exception to this procedure was when participants met the criteria for both statin-induced myopathy case and statin-induced rhabdomyolysis case: for these instances, the index event was set to the earliest qualifying statin-induced rhabdomyolysis case (i.e., the most severe event type). Cases were organized into mutually exclusive groups between induced myopathy case and statin-induced rhabdomyolysis.

Participants met the criteria as a potential control if they had ≥ 1 statin pharmacy record not linked to CK levels > 5× ULN or rhabdomyolysis. The index statin for each controls was set as the first statin dispensing record with a daily dose ≥ 40 milligrams for a statin type that was included in the study. Among the pool of potential controls, we randomly selected those that matched any of the identified cases (10 controls per case). We used propensity matching to balance cases versus controls based on age, biological sex, statin type, and statin dose at index (via MatchIt R package)(10,11). All cases for which we identified 10 controls as well as their corresponding controls made up the analyzed study population for this study. This approach was used to develop the drug response phenotype for AoURP and GERA. Further details are presented in Supplemental Table 1.

Phenotype (bilirubin levels).

Severe statin-induced myopathy is a rare outcome. Furthermore, our variants of interest are common in AFR groups (>1%), but still have low allele frequencies (<10%). Quantitative outcomes have improved statistical power in detecting a genetic effect over rare dichotomous outcomes(12). Thus, as a secondary outcome to support our primary objective, we used total bilirubin levels, an endogenous biomarker for OATP1B1 activity(13). In addition to inherently providing more power than rare outcomes, total bilirubin is routinely measured as part of the general chemical laboratory panel given to most patients (and was included in the UKB biomarker panel); therefore, many more participants had total bilirubin data available than those who were taking statins. Altogether, although this biomarker is not a drug response phenotype, it provided complimentary insight on the role of our candidate variants in statin-induced myopathy. We used bilirubin data from UKB and AoURP. More information about total bilirubin levels in UKB can be found at this link: https://biobank.ndph.ox.ac.uk/showcase/field.cgi?id=30840.

Cell-based assay studies.

Creation of cell lines expressing OATP1B1 reference, OATP1B1-Val174Ala, and OATP1B1-Gly488Ala. The OATP1B1 reference open reading frame (ORF) was cloned into a landing pad vector containing a BxB1-compatible attB recombination site using BsmBI mediated golden gate cloning(14). Site-directed mutagenesis was performed using a Q5 site-directed mutagenesis kit (Catalog no. E0554S, New England Biolabs) to create two missense variants: OATP1B1-Val174Ala and OATP1B1-Gly488Ala. NEBaseChanger tool was used to design primers. Q5® Site-Directed Mutagenesis Kit was used to perform mutagenesis. The HEK293T landing pad cell lines used for this study are previously described(15). To create HEK293T landing pad cell lines expressing OATP1B1, we co-transfected 1,500 nanograms of a library landing pad vector containing OATP1B1 reference, OATP1B1-Val174Ala, or OATP1B1-Gly488Ala with 1,500 nanograms of a BxB1 expression construct (pCAG-NLS-BxB1) using 10.5 microliters of lipofectamine LTX reagent. This transfection was performed in a 6-well plate. All cells were cultured in 1X Dulbecco's Modified Eagle Medium (DMEM, Thermo Fisher Scientific, Inc.), 10% fetal bovine serum (FBS), and 1% penicillin/streptomycin. The HEK293T based cell line has a tetracycline induction cassette upstream of a BxB1 recombination site and split rapamycin analog inducible dimerizable Casp-9. Two days following transfection, expression of integrated genes or iCasp-9 selection system is induced by the addition of doxycycline (2 µg/µL, Sigma-Aldrich) to DMEM-10 media. Two days after induction with doxycycline, AP1903 is added (10 nanomolar, MedChemExpress) to cause dimerization of Casp9. Successful recombination shifts iCasp-9 out of frame, so only non-recombined cells will die from iCasp-9 induced apoptosis following the addition of AP1903. After two days of AP1903-Casp9 selection the media is changed back to DMEM-10 with doxycycline and cells are allowed to recover for two days. After allowing cells to recover for two days, media was changed to DMEM-10 with doxycycline and puromycin (2 microgram per milliliter, Life Technologies Corporation), as an additional selection step to remove non-recombined cells. Cells remained in DMEM-10 plus doxycycline and puromycin for at least two days until cells stopped dying. These cells are then ready for the experiments described below.

Fluorescence-activated cell sorting for abundance assay. After 48-hour transient transfection of mNG2-1-10 expression vector, cells were analyzed using a Becton Dickinson (BD) FACSAria II cell sorter to determine the green fluorescent protein (GFP) expression levels of each cell line. The landing cells expressing OATP1B1 reference, OATP1B1-Val174Ala, OATP1B1-Gly488Ala or no transfection of mNG2-1-10 expression vector were washed, trypsinized, and resuspended in Hank's Balanced Salt Solution (HBSS, Gibco, #14025092) containing 5% FBS. During the mNeongreen and mCherry fluorescence was excited with a 488 nanometer laser and recorded with a 530/30 nanometer band pass filter and 561 nanometer laser and 585/42 nm band pass filter and gated based once mNeonGreen fluorescence doesn't depend on mCherry fluorescence. The GFP/mCherry ratio for each cell was quantified using the FACSAria software.

Cellular methotrexate-fluorescein uptake. The accumulation of methotrexate-fluorescein molecules was measured. The HEK293T landing cells stably expressing OATP1B1 reference, OATP1B1-Val174Ala, OATP1B1-Gly488Ala or no transporter plated onto 6-well plate containing doxycycline to induce the transporter the day before the uptake. On the day of the experiment, the reaction was initiated with the addition of HBSS containing 1 micromolar of methotrexate fluorescein. Uptake was stopped after 20 minutes of incubation at 37°C by removing the HBSS buffer and washing twice with cold HBSS containing 1 micromolar cyclosporin A to stop the reaction. The cells were trypsinized and resuspended in HBSS containing 5% FBS and 1 micromolar cyclosporin A. The cellular fluorescence of the cells was determined using FACSAria II at an excitation wavelength of 488 nanometers and an emission wavelength of 530/30 nanometers. Each variant was assayed in two biological replicates.

Fluorescence microscopy. For the immunostaining experiments, HEK293T landing pad cells stably transfected with OATP1B1 reference, OATP1B1-Val174Ala or OATP1B1-Gly488Ala cell lines were plated onto poly-D-lysine treated 24-well plates at a density of 100,000 cells per well. One day post-seeding, when cells reached 70-80% confluency, they were transiently transfected with mNG2-1-10 expression vector. Two days following transfection, the cell media was removed, and cells were washed with cold phosphate-buffered saline (PBS, Thermo Fisher Scientific, Inc.). The plasma membrane was stained first with Wheat Germ Agglutinin (WGA) Alexa Fluor 647 conjugate (Invitrogen Life Sciences Corporation), diluted in PBS at 1:500, for 15 minutes at room temperature (RT). After staining, the solution was removed, and cells were washed three times with HBSS. Cells were fixed with 3.7% formaldehyde in HBSS for 20 minutes, and after aspiration, cells were washed again three times with HBSS. The nucleus was then stained with Hoechst solution (Thermo Fisher Scientific Inc.), diluted at 1:2000 in HBSS, for 20 minutes at RT in darkness. After staining, the solution was aspirated, and cells were washed twice with HBSS. Coverslips were carefully mounted on Superfrost Plus Microscope Slides (Thermo Fisher Scientific Inc.) with a drop of SlowFade Gold Antifade mountant (Thermo Fisher Scientific Inc.). Slides were left to dry overnight in darkness, and then imaged on an inverted Nikon Ti microscope equipped with a CSU-22 spinning disk confocal. All images were captured with the following channel settings; DAPI at 300ms exposure time and 50% laser power, FITC at 300ms exposure time and 25% laser power, and CY5 at 100ms exposure time and 5% laser power. The images were overlapped using Fiji software.

Transporter assay. The *in vitro* uptake assays were performed using methods developed in our laboratory as previously described(16,17). The HEK293T landing pad cells stably transfected without or with OATP1B1 reference, OATP1B1-Val174Ala or OATP1B1-Gly488Ala were plated in a 48-well plates (poly-d-lysine coated). The culture medium was removed, and cells were washed once with 400 microliters of HBSS. Two radiolabeled substrates of OATP1B1, [$^3$H]-estrone sulfate (Perkin Elmer, NET203250UC) or [$^3$H]-estradiol-17β-glucuronide (American Radiolabeled Chemicals, #ART 1619), were used in the assay. To each well, 150 microliters of HBSS containing trace amounts of radioligand was added to each well and then incubated at 37°C for 15 min. After a 15 minute incubation period, the reaction mix was aspirated, and the cells were washed twice with ice-cold HBSS (800 microliters). Next, 200 microliters of lysis buffer (0.1 normal NaOH, 0.1% SDS) were added to each well and the plate was shaken at room temperature for 60 to 90 minutes. Radioactivity in each well was measured on a Beckman Scintillation counter. To determine function, each variant was normalized to HEK293T landing pad cells without transporter. Each variant was measured in two biological replicates and assayed in triplicate wells.

Cytotoxicity assay. Cytotoxicity of rosuvastatin, was adapted from a previous study(18). We determined the inhibition potency of rosuvastatin to reduce cell count by 50% ($IC_{50}$). First, we seeded cells on a 96-well plate (poly-d-lysine coated) at density 4000 cells per well. After 16-24 hours, the cells were incubated with different concentrations of rosuvastatin, graded from 500 micromolar to 0.7 micromolar, for 72 hours. Doxycycline (2 micrograms per milliliter) was incubated in the media with rosuvastatin to induce the expression levels of OATP1B1. After 72 hours, media was removed, and 50 microliters of media was added to each well. Then 50 microliters of Promega® CellTiter-Glo® luminescent reagent was added to each well. After 10-15 min incubation, we transferred 80 microliters of each well's content to a 96 well plate (white, opaque). The luminescent cell viability was read on the Promega plate reader. The assay was based on quantification of the adenosine triphosphate (ATP) present in each well, which relates to the number of viable cells in each well. The luminescent signal from cells treated with DMSO alone was considered the maximal signal (i.e. 100% cell growth). The percent cell growth of each OATP1B1 reference and mutants and at each concentration of rosuvastatin were calculated. $IC_{50}$ were determined using GraphPad (Prism v9.0).

**Tables**

Supplemental Table 1. Slight variations in methods for the statin-induced myopathy + rhabdomyolysis genetic association study (primary analysis) by cohort

| Method | AoURP | GERA |
|---|---|---|
| Propensity matching using MatchIt R package(10,11)1 | Used “optimal” string values within the *method* argument | Used “optimal” string values (continues variables: age) and “exact” string values (categorical variables: age, biological sex, statin type, statin dose) within *method* argument |
| Age requirement | None | Cases must have experienced a statin-induced myotoxicity event before the age of 91, consistent with previous GERA studies(19) to protect participant privacy |
| Drug interaction | 0.5 years was set as the threshold for an interacting drug to disqualify a statin-induced myotoxicity event (i.e., interacting drug window of use must be within 0.5 years prior to the event) | 1 year was set as the threshold for an interacting drug to disqualify a statin-induced myotoxicity event |

1Matching was performed to balance case versus controls based on age, sex at birth, statin type, and statin dose at index

AoURP, All of Us Research Program; GERA, Genetic Epidemiology Research on Adult Health and Aging

Supplemental Table 2. Characteristics of the statin-induced myopathy + rhabdomyolysis study population at time of index

| Cohort | Race1 | Characteristics | | Case | Control |
|---|---|---|---|---|---|
| AoURP | Black | Total sample size | | 31 | 310 |
| | | Age, median year (IQR) | | 58 (54-66) | 58 (54-65) |
| | | Female sex, n (%) | | 12 (39%) | 120 (39%) |
| | | Statin type/dose, n | | | |
| | | | atorvastatin 40 | 14 | 140 |
| | | | atorvastatin 80 | 9 | 90 |
| | | | fluvastatin 40 | 1 | 10 |
| | | | pravastatin 40 | 3 | 30 |
| | | | pravastatin 80 | 1 | 10 |
| | | | simvastatin 40 | 3 | 30 |
| | White | Total sample size | | 38 | 380 |
| | | Age, median year (IQR) | | 61 (51-66) | 62 (55-69) |
| | | Female sex, n (%) | | 17 (45%) | 170 (45%) |
| | | Statin type/dose, n | | | |
| | | | atorvastatin 40 | 16 | 160 |
| | | | atorvastatin 80 | 10 | 100 |
| | | | fluvastatin 40 | 4 | 40 |
| | | | pravastatin 40 | 1 | 10 |
| | | | pravastatin 80 | 5 | 50 |
| | | | simvastatin 40 | 2 | 20 |
| GERA | Black | Total sample size | | 40 | 392 |
| | | Age, median year (IQR) | | 67 (61-73) | 67 (63-72) |
| | | Female sex, n (%) | | 13 (33%) | 130 (33%) |
| | | Statin type/dose, n | | | |
| | | | atorvastatin 40 | 8 | 80 |
| | | | lovastatin 40 | 8 | 80 |
| | | | pravastatin 40 | 2 | 20 |
| | | | simvastatin 40 | 7 | 70 |
| | | | atorvastatin 60 | 1 | 2 |
| | | | atorvastatin 80 | 4 | 40 |
| | | | lovastatin 80 | 2 | 20 |
| | | | pravastatin 80 | 1 | 10 |
| | | | simvastatin 80 | 7 | 70 |
| | White | Total sample size | | 446 | 4460 |
| | | Age, median year (IQR) | | 77 (69.7-82.9) | 77 (69.7-82.8) |
| | | Female sex, n (%) | | 205 (46%) | 2,050 (46%) |
| | | Statin type/dose, n | | | |
| | | | atorvastatin 40 | 97 | 970 |
| | | | lovastatin 40 | 93 | 930 |
| | | | pravastatin 40 | 8 | 80 |
| | | | rosuvastatin 40 | 3 | 30 |
| | | | simvastatin 40 | 80 | 800 |

| | | | | | |
|---|---|---|---|---|---|
| | | | atorvastatin 60 | 1 | 10 |
| | | | lovastatin 60 | 6 | 60 |
| | | | simvastatin 60 | 3 | 30 |
| | | | atorvastatin 80 | 33 | 330 |
| | | | lovastatin 80 | 42 | 420 |
| | | | pravastatin 80 | 11 | 110 |
| | | | simvastatin 80 | 69 | 690 |

1Self-identified race

AoURP, All of Us Research Program; GERA, Genetic Epidemiology Research on Adult Health and Aging; IQR, interquartile range.

Supplemental Table 3. Characteristics of the total bilirubin study population in self-identified Black participants at time of index

| Characteristics | UKB (N=6,651) | AoURP (N=22,468) |
|---|---|---|
| Female sex | 3,744 (66%) | 14,813 (66%) |
| Age, mean year (SD) | 52.5 (8.1) | 52.1 (14.9%) |
| Body mass index1, mean kilograms per meter squared (SD) | 29.5 (5.4) | 31.7 (7.2) |
| Total bilirubin12 | 7.8 (6.0-10.5) | Mean 0.44 (SD 0.19) |
| Female total bilirubin levels | 7.0 (5.5-9.2) | Mean 0.41 (SD 0.18) |
| Male total bilirubin levels | 8.9 (6.9-12.1) | Mean 0.50 (SD 0.18) |

1Body mass index and total bilirubin levels are from the first assessment visit per the cohort study design (UKB) or the first chronological measurement available in the dataset (AoURP). Total bilirubin levels are in the units of micromoles per liter (UKB) or milligrams per deciliter (AoURP).

AoURP, All of Us Research Program; SD, standard deviation; UKB, UK Biobank.

Data presented as median (interquartile range) or count (%) unless indicated otherwise.

Supplemental Table 4. The relationship1 between *SLCO1B1* variants and total bilirubin levels in self-identified Black participants from AoURP (N=22,468)

| *SLCO1B1* variant | Effect allele frequency | Beta1 | SE | P-value |
|---|---|---|---|---|
| c.481+1G>T | 0.030 | 0.038 | 0.005 | $2.5x10^{-14}$ |
| c.1463G>C | 0.034 | 0.026 | 0.005 | $2.9x10^{-8}$ |
| c.521T>C | 0.033 | 0.023 | 0.005 | $1.8x10^{-6}$ |

1Generated by linear regression of log-transformed total bilirubin levels. Covariates included age, sex at birth, body mass index at the time of measurement, and the first 10 principal components of population-specific genetic similarity.

AoURP, All of Us Research Program

Supplemental Table 5. The relationship1 between *SLCO1B1* variants and total bilirubin levels in self-identified Black participants with levels within the range of normal2

| | *SLCO1B1* variant | Effect allele frequency | Beta | SE | P-value |
|---|---|---|---|---|---|
| UKB (n=6,461) | c.481+1G>T | 0.027 | 0.100 | 0.020 | $2.04 \times 10^{-7}$ |
| | c.1463G>C | 0.043 | 0.063 | 0.016 | $6.80 \times 10^{-5}$ |
| | c.521T>C | 0.024 | 0.051 | 0.021 | 0.015 |
| AoURP (n=21,854) | c.481+1G>T | 0.030 | 0.039 | 0.006 | $4.82 \times 10^{-11}$ |
| | c.1463G>C | 0.034 | 0.024 | 0.006 | $1.36 \times 10^{-5}$ |
| | c.521T>C | 0.033 | 0.031 | 0.006 | $3.04 \times 10^{-8}$ |

1Generated by linear regression of log-transformed total bilirubin levels. Covariates included age, sex at birth, body mass index at the time of measurement, and the first 10 principal components of population-specific genetic similarity.

2The range of normal was 1.71-20.50 micromoles per liter (UKB) and 0.1-1.2 milligrams per deciliter (AoURP)

AoURP, All of Us Research Program; sd, standard deviation; UKB, UK Biobank.

3. National Academies of Sciences, Engineering, and Medicine; Division of Behavioral and Social Sciences and Education; Health and Medicine Division; Committee on Population; Board on Health Sciences Policy; Committee on the Use of Race, Ethnicity, and Ancestry as Population Descriptors in Genomics Research. Using population descriptors in genetics and genomics research: A new framework for an evolving field. Washington (DC): National Academies Press (US); 2023.

9. Oni-Orisan A, Lu M, Peng JA, Krauss RM, Iribarren C, Medina MW. Development and application of an algorithm for statin-induced myopathy based on electronic health record-derived structured elements. medRxiv. 2023 Apr 25;
